# Importance of arterial measurement sites for intraoperative hemodynamic management during major abdominal surgeries: The IPAMS randomized trial protocol

**DOI:** 10.1101/2025.11.02.25339344

**Authors:** Shi Shu Wang, Mathias Pietrancosta, Louis Morisson, Hugo Langlois, Thomas Hemmerling, Olivier Verdonck, Philippe Richebé, André Denault, Pascal Laferrière-Langlois

**Affiliations:** Maisonneuve-Rosemont Hospital Research Center, Montreal, Quebec, Canada; Faculty of Medicine, University of Montreal, Montreal, Quebec, Canada; Department of Anesthesiology and Pain Medicine, Maisonneuve-Rosemont Hospital, Montreal, CIUSSS de l’Est de l’Île de Montréal, Montreal, Quebec, Canada; Department of Anesthesiology and Pain Medicine, University of Montreal, Montreal, Quebec, Canada; Department of Anesthesiology, Montreal Heart Institute, University of Montreal, Montreal, Quebec, Canada; Department of Experimental Surgery, McGill University Health Center, Montreal, Quebec, Canada; Department of Anesthesia, McGill University, Montreal, Quebec, Canada; Polyclinique Bordeaux Nord Aquitaine, Bordeaux, France

## Abstract

**Introduction:** Reliable hemodynamic parameters are crucial to prevent perioperative hypotension, one of the most significant modifiable risk factors for postoperative morbidity and mortality. The current standard of care for intraoperative hemodynamic monitoring is achieved through an arterial radial catheter, but its reliability has been increasingly questioned due to the incidence of a phenomenon called the radial-to-central arterial pressure gradient. Studies have shown that this gradient occurs in one-third of cardiac surgery patients predominantly during moments of hemodynamic instability, when accurate blood pressure monitoring is critical. However, it remains unclear whether and how this phenomenon affects non-cardiac surgical populations.

**Objectives:** The main objective is to study whether brachial arterial pressure monitoring reduces intraoperative vasopressor dosage compared to conventional radial monitoring. We hypothesized that patients undergoing major abdominal surgery with brachial arterial pressure monitoring will require lower time-weighted average of phenylephrine administration compared to those monitored with radial arterial pressure. Our secondary objectives are to systematically describe the incidence of radial-to-brachial pressure gradients and identify predictive factors associated with this phenomenon.

**Materials and methods:** This single center randomized clinical trial will enroll 200 adult patients between 18 and 90 years of age, undergoing major abdominal surgery. Besides standardized anesthesia management, all patients will receive radial and brachial blood pressure monitoring, but only one blinded catheter will guide intraoperative hemodynamic management. We will record radial and brachial arterial pressure curves, standard intraoperative data and post-operative complications.

**Ethics:** This trial has been approved by the regional ethics committee (Comité d’Éthique de la Recherche du CIUSSS de l’Est de l’Île de Montréal)

**Trial registration:** https://ClinicalTrials.gov (May 21st, 2025). Unique protocol ID: 2023-3299. Trial identification number: NCT06982001

## 1. Introduction

### 1.1 Background and rationale

Perioperative hypotension represents one of the most significant modifiable risk factors for postoperative morbidity and mortality (1, 2). The Canadian Anesthesiologists’ Society and American Society of Anesthesiologists mandate continuous invasive blood pressure monitoring during general anesthesia when hemodynamic risk is elevated (3, 4).

However, growing evidence from cardiac surgery populations questions the reliability of radial artery pressure monitoring, the current standard of care. Studies demonstrate that significant radial-to-central pressure gradients (i.e. measured on brachial, femoral) occur in approximately one-third of cardiac surgery patients, with gradients present during 25% of operative time and more frequently in patients with radial artery diameter less than 1.8 mm (5, 6). These gradients predominantly occur during hemodynamic instability when accurate monitoring is most critical, and patients monitored exclusively with radial arterial lines demonstrate increased vasopressor requirements and prolonged vasoactive medication use compared to those with dual monitoring approaches (5, 7). The clinical implications of inaccurate arterial pressure monitoring are profound. Both depth and duration of perioperative hypotension correlate strongly with postoperative mortality and organ dysfunction, with extensive literature linking inadequate blood pressure control to myocardial infarction, acute kidney injury, and stroke (8–10). Conversely, excessive vasopressor administration carries risks including myocardial strain, arrhythmias, compromised microcirculation, and renal failure. Recent precision medicine approaches demonstrate that maintaining blood pressure within 20% of patient baseline reduces postoperative organ dysfunction compared to fixed thresholds, emphasizing the importance of accurate pressure measurement (11). While radial-to-central pressure gradients are well-characterized in cardiac surgery, no studies have systematically investigated this phenomenon in major abdominal surgery populations. This represents a significant knowledge gap given that major abdominal surgery often requires intensive hemodynamic monitoring due to fluid shifts, blood loss, and surgical stress. Despite not being exposed to cardiopulmonary bypass, major abdominal surgery can involve vena cava compression with significant reduction in cardiac preload, aortic clamping for aortic repair, hyperthermic chemotherapy during oncological surgery, extreme Trendelenburg position (head down), as well as many other conditions. The brachial artery presents a theoretically superior monitoring site due to its larger diameter (3.19±0.75 mm versus 1.77±0.48 mm for radial arteries) and more central location, with recent evidence demonstrating acceptable safety profiles with minimal complications (12, 13). Canada performs over two million surgeries annually, with a substantial proportion requiring invasive arterial monitoring (14). The current evidence gap regarding optimal monitoring site selection in noncardiac surgery represents both a patient safety concern and an opportunity for evidence-based practice improvement, not only in major tertiary care centers, but in in all hospital providing surgeries (15). Therefore, it is essential to compare brachial arterial monitoring with the current standard of radial monitoring to determine whether the use of a more central site is justified in specific surgical contexts. Demonstrating the superiority of brachial over radial monitoring or identifying patient characteristics in which brachial monitoring provides distinct advantages, would offer clinicians evidence-based guidance on optimal site selection, and inform future practice guidelines through a patient-specific approach to arterial pressure monitoring.

### 1.2 Objectives and hypothesis

The primary objective of this study is to determine whether brachial arterial pressure monitoring impacts hemodynamic guidance management when compared to radial monitoring during major abdominal surgery. We hypothesize that brachial arterial pressure monitoring reduces vasopressor administration compared to the current standard of radial monitoring.

Our secondary objectives included the following:

– Systematically describe the impact of monitoring site on fluid administration.
– Systematically describe the context of appearance of the radial-to-brachial pressure gradients.
– Identify patient and procedural factors that are associated with the occurrence of this phenomenon.
– Evaluate postoperative cardiac and renal status through biomarkers.
– Evaluate postoperative complications through the Clavien-Dindo Classification system

### 1.3 Trial design

This is the protocol for a prospective, single-center, parallel-group, double-blinded randomized trial with two intervention arms.

## 2. Methods

This protocol follows the SPIRIT 2025 guidelines for randomized trial protocols (16). Our SPIRIT 2025 checklist is presented in supplementary information file (S1). The original protocol submitted to the ethics board can be found in supplementary information file (S2).

### 2.1 Participants

This trial will be conducted at the Maisonneuve-Rosemont Hospital which is a part of the Centre intégré universitaire de santé et de services sociaux (CIUSSS) de l’est de l’Île-de-Montréal (CEMTL), located in Montreal, Quebec, Canada. Patients will be included in our study if they meet the following inclusion and exclusion criteria.

Inclusion criteria:

– Adult patients between 18 to 90 years of age undergoing major abdominal surgery (general, vascular, gynecological, urological) via laparoscopy or laparotomy with expected anesthesia duration of at least 120 minutes.
– Clinical indication for invasive arterial pressure monitoring during surgery
– Surgical position that allows bilateral arterial catheterization without compromising clinical care
– Ability to provide informed consent and communicate in French or English

Exclusion Criteria:

– Known severe peripheral vascular disease
– Significant preoperative inter-arm pressure gradient, defined as a difference greater than 5 mmHg in mean arterial pressure
– Contraindications to bilateral arterial line placement (i.e., arteriovenous fistula)
– Known thrombophilia with increased thrombosis risk

### 2.2. Interventions

Following informed consent, baseline vital signs will be recorded including bilateral non-invasive blood pressure measurements to screen for the exclusion criteria of significant inter-arm gradients. Patients will undergo comprehensive preoperative assessment including demographic data collection, medical history review, and physical examination. Upon entry in the operating room, standard anesthesia preparation will be made, including establishment of peripheral intravenous access, and application of standard monitoring equipment. A first arterial line can be inserted prior to general anesthesia induction. The second arterial line will be inserted after general anesthesia induction to minimize patient’s discomfort. If no arterial line was inserted prior to anesthesia induction, both lines will be inserted under general anesthesia. Study protocol and outcome assessment will start once both arterial lines are positioned, that patient positioning if finalized, proper blinding has been initiated, and surgical incision is being made. All arterial lines will be installed using ultrasound-guidance and with 20-gauge catheters. Prior to insertion, vessel diameter, vascular and blood velocity will be documented for subsequent analysis. Both arterial lines are connected to pressure transducers and zeroed at the level of the right atrium, with simultaneous pressure recording initiated and maintained throughout the procedure. Electronic blinding will be implemented through software modification preventing display of the non-assigned arterial line to clinical staff while maintaining complete data recording. Anesthesia induction will follow a standardized protocol including pre-oxygenation, intravenous lidocaine 40mg, propofol 1.5mg.kg^-1^, remifentanil 1μg.kg^-1^ over 30 seconds, and rocuronium 0.8mg.kg^-1^ for neuromuscular blockade. Endotracheal intubation is performed when appropriate neuromuscular blockade depth is achieved, followed by mechanical ventilation using volume-controlled mode with positive end-expiratory pressure of 5 cmH_2_O respiratory rate of 14 breaths per minute, and tidal volumes adjusted to maintain end-tidal CO2 between 35-45 mmHg. Analgesic administration during anesthesia maintenance will be standardized based on nociception level. Anesthesia will be maintained to achieve bispectral index (BIS) values between 40 and 60, using either propofol or sevoflurane, depending on the anesthesiologist’s preference. Standardized protocols with baseline mean arterial pressure defined as the average of three consecutive measurements taken one minute apart before anesthesia induction, and phenylephrine infusion initiated at 0.2 μg.kg^-1^.min^-1^ and titrated to maintain mean arterial pressure within 20% of baseline values. The algorithm for anesthesia maintenance can be found under supplementary material (S3 File). Fluid management will be guided by pulse pressure variation measurements obtained every 30 minutes, with crystalloid boluses of 250-500 mL administered when pulse pressure variation exceeds 12%. All medication administration, hemodynamic interventions, and significant clinical events are timestamped and recorded in the electronic data collection system. Using the timestamps for phenylephrine dose change, we will reconstruct the administration profile for the whole surgery duration. Every 30 minutes, we will record the total dose of phenylephrine administered as a validation, the total fluids administered, and the urine output. All data from the anesthesia workstation as well as the BIS and NOL index will be extracted on a research computer. Upon completion of surgery, anesthesia agents will be discontinued, and neuromuscular blockade will be reversed using appropriate reversal agents when indicated. Patients will receive standardized postoperative medications and extubation will be performed in the operating room when standard criteria will be met. Both arterial lines will be removed in the post-anesthesia care unit unless transferred to the intensive care unit. We will record any complications related to arterial lines and perform ultrasound assessment if needed. An assessment of the neurovascular functions of the upper limbs will be performed postoperatively to evaluate any complications due to arterial line placement. Postoperative pain management will follow institutional enhanced recovery after surgery protocols. In post-anesthesia care unit prior to discharge and post-operative day 1,3 and 7, we will collect troponin and creatinine (test labs for heart and kidney function) and assess for postoperative complications using the Clavien-Dindo classification.

### 2.3 Outcomes

Our primary outcome is the time-weighted average (TWA) phenylephrine administration (μg.kg^-1^.min^-1^) from surgical incision to wound closure, calculated as total phenylephrine dose divided by procedure duration, normalized for patient weight.

Our secondary outcomes are categorized into hemodynamic, safety and mechanistic domains, and include the following:

Hemodynamic outcome:

– total phenylephrine dose administered
– time below hypotension threshold (baseline MAP -20%)
– radial-to-brachial pressure gradient incidence and magnitude
– fluid balance indicators

Safety outcomes:

– arterial line-related complications
– postoperative organ dysfunction markers (troponin, creatinine)
– hospital length of stay, and 30-day complications using Clavien-Dindo classification

Mechanistic outcomes:

– radial artery diameter correlations with gradient occurrence
– temporal patterns of pressure gradient development

### 2.4 Recruitment and participant timeline

Our site hosts between 10-15 eligible surgeries weekly, creating a large pool of eligible patients and ensuring adequate recruitment rates for timely study completion. With a conservative inclusion rate of three to five participants per week, and with monthly monitoring of recruitment to ensure adequacy and allow adjustments if needed, we are confident that our target sample size will be reached within the planned two-year study period.

By consulting our hospital’s surgical schedule, the research team will screen for eligible patients one week prior to their surgery using the institution’s electronic medical records. Eligible patients will then be approached during their preoperative clinic (CIEPC) visit or, alternatively, via a phone call 48 hours prior to surgery to assess their interest in participation. On the day of surgery, participants will be reassessed in the preoperative ward, where they will be reinformed of the study details and provided with the consent form. Participants will be encouraged to take time to review the form, and all questions will be addressed. Final enrollment will be confirmed upon signing the consent form on the day of surgery, after which patients will be randomized to either the brachial-guided group or the radial-guided group. Postoperative assessments will be conducted in the post-anesthesia care unit and on postoperative days one, three, and seven. For participants discharged before day seven, follow-up assessments will be performed by telephone. A completed SPIRIT schedule of enrollment, interventions and assessments is presented in Fig 1. (17)

**Fig 1.**
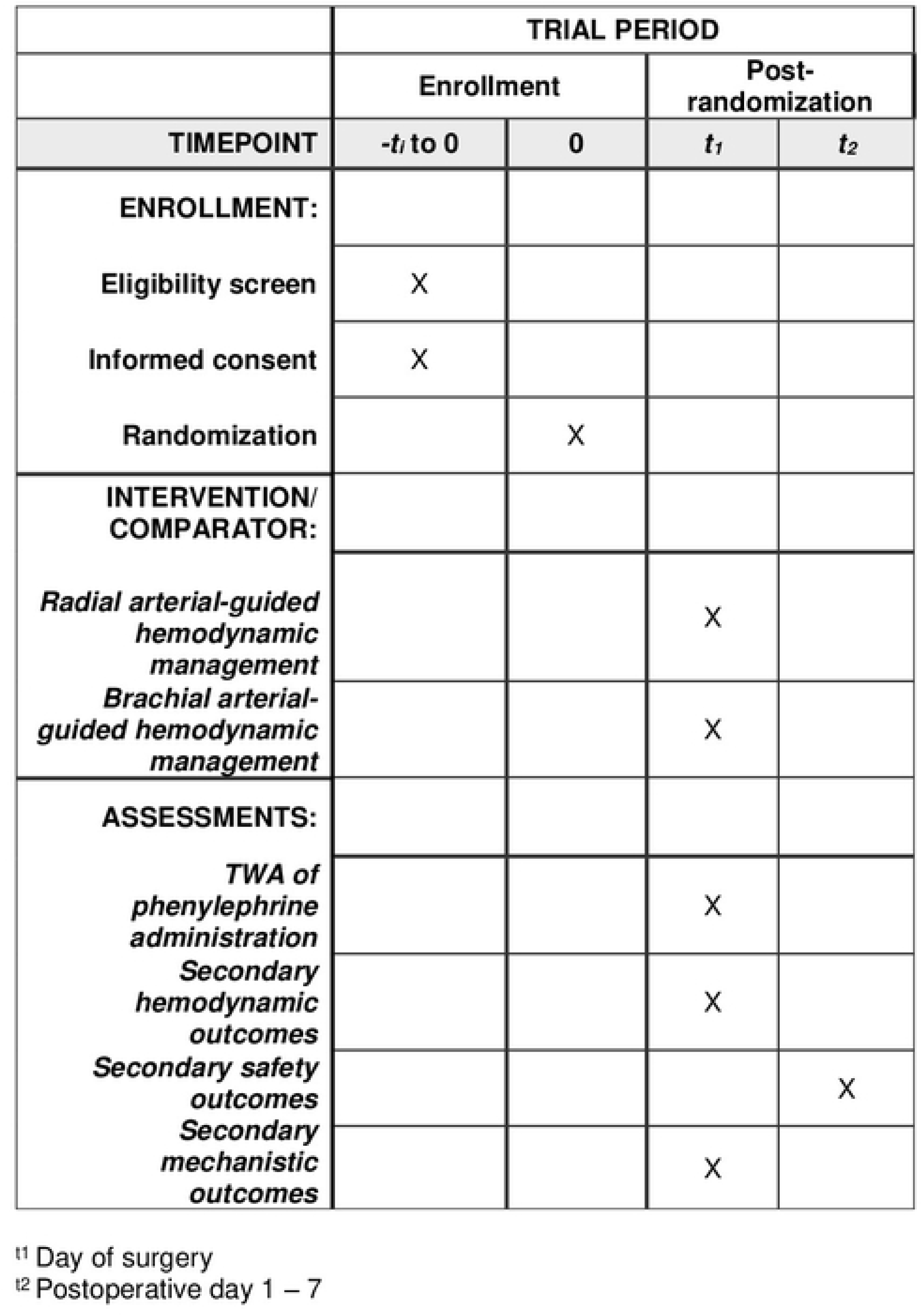
SPIRIT Schedule of enrollment, interventions, and assessments

### 2.5 Sample size and recruitment

Sample size calculations are based on the primary outcome of time-weighted average phenylephrine administration, utilizing data from previous perioperative studies conducted at our center, and institutional experience with major abdominal surgery. Literature reports from similar surgical populations indicate mean phenylephrine requirements of 0.35±0.25 μg.kg^-1^.min^-1^ during procedures lasting more than 2 hours. Based on preliminary data and published reports of clinical differences between monitoring sites, we conservatively estimate that brachial monitoring could reduce vasopressor requirements by 30% compared to radial monitoring, representing a clinically meaningful difference of approximately 0.105 μg.kg^-1^.min^-1^. Using a two-sample t-test with equal variances, achieving 80% statistical power to detect this difference with a two-sided alpha level of 0.05 requires 88 patients per group. To account for potential protocol violations, arterial line failures, and data quality issues, we will recruit 200 patients (100 per group), providing 10% inflation above the minimum required sample size. We anticipate starting patient recruitment on October 1^st^, 2025, until December 31^st^, 2027. We expect data collection to be completed and report writing to begin in Q4 of 2027. Following analysis, we plan on presenting our results to conferences by Q4 of 2028.

### 2.6 Randomization and blinding

Participants will be randomized in a 1:1 ratio to the monitoring groups using computer-generated allocation. The research personnel responsible for participant enrollment and assignment will remain blinded to the random allocation sequence. Group assignments will be concealed in opaque envelopes, which will be opened by study personnel only after completion of arterial line placement. All patients will receive bilateral arterial catheters (radial and contralateral brachial) with simultaneous high-fidelity pressure recording and data extraction. The assigned group will determine which arterial line guides hemodynamic management while the alternate line will remain electronically blinded to clinical staff. Both the staff and the patient will be blinded to the arterial line use to guide for hemodynamic management. To maintain blinding during surgery, once installed, the two arterial lines will go through an opaque tube in such a way that the attending clinician does not know which line is which. In case of any manipulations of a line (i.e. flushing), both lines will be flushed. In case of any required manipulations under the drape (repositioning of the lines), a dedicated research staff will be responsible of performing the manipulation. This blinding strategy has undergone multiple tests and has been demonstrated effective at maintaining blinding.

### 2.7 Data collection and management

Data for primary and secondary outcomes will be collected at baseline, intraoperatively and postoperatively. Participant socio-demographic characteristics, medical history and current medication will be extracted from medical records, while postoperative data will be obtained from medical records or through telephone follow-up. Intraoperatively, all electronic data from the medical monitoring systems including BIS^TM^ and PMD-200^TM^ will also be collected. Monitor times will be synchronized before any data is collected. By using a third-party software named Bettercare, we will extract the high-fidelity waveforms and perioperative parameters. We will note on a separate Case Report Form (CRF) and in the integrated system of PMD-200^TM^, all anesthesia and surgery related events. At the end of anesthesia, and once the participant is extubated, we will export all electronic data.

To extract the high-frequency arterial waveforms, we will extract data from the Dräger anesthesia monitors by using an application programming interface (API) and a physical connection to our research computers, combined with the Bettercare software (Bettercare inc., Spain). With this solution, we extract physical waveform at 200 Hz, as well as other parameters monitored during anesthesia (EKG, capnography, etc.)

Hard copies of patient consent forms will be securely stored in a locked file cabinet within the offices of the Department of Anesthesiology and Pain Medicine at Maisonneuve-Rosemont Hospital, Montreal, Quebec, Canada. Electronic data will be maintained on a password-protected laptop assigned to Dr. Pascal Laferrière-Langlois, who will have primary responsibility for both the device and the data it contains. Each participant will be assigned a unique study code to link their data to their identity. This code key will be stored separately to prevent participant identification in the event of theft or loss. During the study, only the principal investigator will have full access to all participant data.

### 2.8 Statistical Analysis

Descriptive statistics will be presented by study group. Continuous variables will be summarized using mean (standard deviation) or median (interquartile range), depending on the distribution of the data, while categorical variables will be described using frequencies and percentages. Differences between study groups, according to the type of endpoint, will be reported with 95% confidence intervals for proportions or means/medians as appropriate. A two-sided significance level of 0.05 will be used, and all statistical analyses will be conducted using SAS, SPSS, R, or Python

The primary analysis will follow the intention-to-treat principle and assess the difference between brachial and radial arterial monitoring in the intraoperative time-weighted average dose of phenylephrine administered. Depending on the distribution of the variable, this comparison will be performed using a t-test or a Mann-Whitney-Wilcoxon test.

Secondary continuous outcomes will be analyzed using the same approach as the primary outcome. Dichotomous secondary endpoints will be summarized using frequencies and percentages by randomized group, and comparisons between groups will be conducted using the chi-square test or Fisher’s exact test, as appropriate. Risk differences or mean differences with 95% confidence intervals will be reported for all outcomes.

Given that not all participants are expected to remain hospitalized until postoperative day 7, some missing data are anticipated, particularly for laboratory results. For patients discharged before day 7, we will assume normal laboratory values in the absence of readmission. For patients readmitted with complications, laboratory results from the readmission will be extracted for the corresponding postoperative day. While this approach represents an estimate rather than the true values, the expected low rate of organ dysfunction suggests minimal impact on study findings.

Longitudinal data collected on postoperative days 1, 3, and 7, including laboratory results and Clavien-Dindo scores, will be analyzed using a two-way repeated measures ANOVA with factors for study treatment group, timepoint, and the interaction between treatment group and time.

Subgroup analyses of the primary endpoint will be performed according to the following categories: surgical type (vascular, general, or gynecological), procedure duration (≤4 hours vs. >4 hours), radial artery diameter (<1.8 mm), participant age (>70 years), and presence of diagnosed vascular disease. Treatment effects (difference in mean or in proportion) will be reported for our primary endpoint with 95% confidence intervals within each subgroup. The treatment effect within each subgroup will be evaluated using a two-way ANOVA, incorporating terms for the study treatment group (brachial artery versus standard of care), the subgroup factor, and the interaction between treatment and subgroup. If the primary endpoint exhibits a non-normal distribution, a log-transformation or an appropriate non-parametric method can be applied

### 2.9 Data monitoring

This study involves minimal additional risk and is not expected to prolong anesthesia beyond approximately 10 minutes required for insertion of a second arterial line, in the context of surgeries lasting over 120 minutes. Both radial and brachial arterial lines are standard techniques for intraoperative hemodynamic management, and anesthetic care will otherwise follow routine practice with standard monitoring and commonly used medications. In light of the low-risk profile and absence of anticipated safety concerns, a formal data monitoring committee, stopping guidelines, or independent trial monitoring are not deemed necessary. The principal investigator will have access to interim results and will assume responsibility for acting upon them if required.

### 2.10 Dissemination policy

Publication of our results will be proposed to The Journal of American Medical Association (JAMA) or The Lancet. Alternatively, our manuscript will be submitted to either the British Journal of Anaesthesia, Canadian Journal of Anesthesia or Anesthesiology. Research staffs that have contributed significantly to the completion of this study and reporting of results will be considered for authorship. There was no patient or public involvement in the design of this trial.

## 3. Ethics

This protocol has been approved by the regional ethics committee (Comité d’éthique en recherche du CIUSSS de l’Est de l’Île de Montréal). Any modifications to the study protocol will be submitted to the regional ethics committee, updated on ClinicalTrials.gov, and communicated to participants or staff when relevant, with all changes documented in the final report. In the event of study termination, participants will be informed, and data handled according to ethical standards. Participants interest in study enrollment will occur either through an initial phone call or during the patient’s preoperative clinic visit prior to the day of surgery. On the day of surgery, participants will be approached by a research staff in the preoperative ward to address any remaining questions, and if they agree to participate, written informed consent will be obtained.

Participant confidentiality will be strictly maintained, with secure storage of all data. Data will only be disclosed when required by law, authorized oversight, or participant permission. All study personnel will be trained in confidentiality procedures in compliance with regulations. Finally, participant safety will remain a priority. Bilateral blood pressure measurements performed prior to inclusion may identify previously undiagnosed vascular disease. Depending on severity, participants may be excluded according to study criteria or remain enrolled. In such cases, a consultation with the appropriate specialist will be requested to ensure appropriate clinical management. In the rare event of an unexpected serious adverse event, the principal investigator, Dr. Pascal Laferrière-Langlois, will be promptly notified and appropriate care provided.

## 4. Discussion

Continuous invasive blood pressure monitoring is critical for hemodynamic management during high-risk surgeries. Therefore, accurate and reliable blood pressure measurements are essential for guiding clinicians in the appropriate administration of vasopressors and fluids, particularly during episodes of hemodynamic instability, when radial-to-central arterial pressure gradients are more likely to occur (6). Our study seeks to address the current knowledge gap regarding the effect of radial-to-central arterial pressure gradients on hemodynamic monitoring among non-cardiac surgical populations. By characterizing the frequency and determinants of this phenomenon, our methodology aims to provide clinicians with an evidence-based alternative for continuous blood pressure monitoring, thereby challenging the prevailing reliance on radial artery measurements alone.

Strengths of the study include the involvement of a highly experienced multidisciplinary team, including collaborators with extensive expertise on intraoperative hemodynamic monitoring and radial-to-central arterial pressure gradients. An additional strength is our open-science approach, with the research protocol published prior to patient enrollment, thereby enhancing transparency and reproducibility. Recruitment feasibility is enhanced by prior expertise in perioperative clinical trials and access to a large pool of eligible participants, ensuring timely enrollment. The study also benefits from specialized equipment for dual-site arterial monitoring, ultrasound-guided catheterization, and advanced hemodynamic monitoring (BIS, NOL), as well as optimized electronic data capture systems. Data collection and analysis are supported by the Research Center of Maisonneuve-Rosemont Hospital, with access to skilled technical personnel who will ensure protocol adherence, data integrity, and continuity of operations. Importantly, the trial is designed as a low-risk, randomized study conducted under a rigorous protocol.

This study design incorporates several inherent limitations that must be acknowledged while considering their impact on result interpretation and generalizability. The single-center design may limit generalizability to other institutions with different practices, though this approach ensures protocol standardization and reduces variability while our institutional volume provides adequate recruitment capacity. Short-term follow-up limitations focus the study on intraoperative outcomes with limited postoperative follow-up, leaving long-term consequences of monitoring strategy differences unexplored and requiring future studies with extended follow-up periods. To assess the occurrence of postoperative organ dysfunction, we monitor the troponin, creatinine and Clavien-Dindo score, but this evaluation is limited to 7 days due to the logistical and cost constraints associated with longer-follow-up. We simultaneously acknowledge that the use of vasopressor administration as a surrogate outcome for clinical decision-making quality may not capture all relevant clinical effects, but this work being one of the first trial exploring this question, some positive findings will path the way to trials exploring for larger trial exploring direct patient-centered outcomes with longer follow-up.

## Data Availability

No datasets were generated or analysed during the current study. All relevant data from this study will be made available upon study completion.

## Supporting information Files

S1 File. SPIRIT 2025 Checklist

S2 File. Original protocol (Approved by Ethics)

S3 File. Anesthesia Maintenance

